# *ANGPTL8* protein-truncating variant and the risk of coronary disease, type 2 diabetes and adverse effects

**DOI:** 10.1101/2020.06.09.20125278

**Authors:** Pyry Helkkula, Tuomo Kiiskinen, Aki S. Havulinna, Juha Karjalainen, Seppo Koskinen, Veikko Salomaa, FinnGen, Mark J. Daly, Aarno Palotie, Ida Surakka, Samuli Ripatti

**Affiliations:** Institute for Molecular Medicine Finland (FIMM), HiLIFE, University of Helsinki, Helsinki, Finland; Finnish Institute for Health and Welfare, Helsinki, Finland; Analytic and Translational Genetics Unit, Massachusetts General Hospital and Harvard Medical School, Boston, MA, USA; Broad Institute of the Massachusetts Institute of Technology and Harvard University, Cambridge, MA, USA; Psychiatric & Neurodevelopmental Genetics Unit, Department of Psychiatry, Analytic and Translational Genetics Unit, Department of Medicine, and the Department of Neurology, Massachusetts General Hospital, Boston, MA, USA; Department of Internal Medicine, University of Michigan, Ann Arbor, MI, USA; Department of Public Health, University of Helsinki, Helsinki, Finland

## Abstract

Protein-truncating variants (PTVs) affecting dyslipidemia risk may point to therapeutic targets for cardiometabolic disease. Our objective was to identify PTVs that associated with both lipid levels and cardiometabolic disease risk and assess their possible associations with risks of other diseases. To achieve this aim, we leveraged the enrichment of PTVs in the Finnish population and tested the association of low-frequency PTVs in 1,209 genes with serum lipid levels in the Finrisk Study (n = 23,435). We then tested which of the lipid-associated PTVs also associated with risks of cardiometabolic diseases or 2,264 disease endpoints curated in the FinnGen Study (n = 176,899). Three PTVs were associated with both lipid levels and the risk of cardiometabolic disease: triglyceride-lowering variants in *ANGPTL8* (−24.0[-30.4 to −16.9] mg/dL per rs760351239-T allele, *P* = 3.4× 10^−9^) and *ANGPTL4* (−14.4[-18.6 to −9.8] mg/dL per rs746226153-G allele, *P* = 4.3 × 10^−9^) and the HDL cholesterol-elevating variant in *LIPG* (10.2[7.5 to 13.0] mg/dL per rs200435657-A allele, *P* = 5.0 × 10^−13^). The risk of type 2 diabetes was lower in carriers of *ANGPTL8* (odds ratio [OR] = 0.67[0.47-0.92], *P* = 0.01), *ANGPTL4* (OR = 0.70[0.60-0.82], *P* = 1.4× 10^−5^) and *LIPG* (OR = 0.67[0.48-0.91], *P* = 0.01) PTVs than in noncarriers. Moreover, the odds of coronary artery disease were 44% lower in carriers of a PTV in *ANGPTL8* (OR = 0.56[0.38-0.83], *P* = 0.004). Finally, the phenome-wide scan of the *ANGPTL8* PTV showed a markedly higher associated risk of esophagitis (585 cases, OR = 174.3[17.7-1715.1], *P* = 9.7 × 10^−6^) and sensorineural hearing loss (12,250 cases, OR = 2.45[1.63-3.68], *P* = 1.8 × 10^−5^). The *ANGPTL8* PTV carriers were less likely to use statin therapy (53,518 cases, OR = 0.53[0.41-0.71], *P* = 1.2 × 10^−5^). Our findings provide genetic evidence of potential long-term efficacy and safety of therapeutic targeting of dyslipidemias.

## Main

Dyslipidemia is a major risk factor for cardiovascular disease and is present in nearly half of type 2 diabetes patients[1]. For treating dyslipidemia, there are few alternatives to low-density lipoprotein (LDL) cholesterol-lowering therapy. Although common, this therapy often fails to treat the condition effectively, leaving patients with high risk of cardiovascular disease[2]. Therefore, a search for possible new drugs is necessary. This could be achieved, firstly, by developing drugs that modulate other blood lipid levels besides LDL cholesterol. Secondly developing new drugs that lower LDL cholesterol levels through different molecular mechanisms than existing ones. Thirdly, maximizing the safety of dyslipidemia drugs to enable their widespread and preventative use.

There is a limited number of drugs for treating dyslipidemia currently on the market or in development. Drugs targeting triglycerides, lipoprotein(a), high-density lipoprotein (HDL) cholesterol that are currently being developed have emerged from studies of PTVs[3-9], and it is uncertain whether they are safe enough to reach the market. Meanwhile, PCSK9 inhibitors and ezetimibe are the only common alternatives to statins for lowering LDL cholesterol levels. More options would be welcome given that statins have common side effects[10], and a combination therapy with ezetimibe does not reduce coronary atherosclerosis compared to patients on statin monotherapy[11]. On the other hand, PCSK9 inhibitors are unaffordable to many patients and do not benefit people with familial hypercholesterolemia, who have a defective copy of *LDLR* that PCSK9 inhibits[12, 13].

Besides the small number of alternatives, another issue is the long-term safety of drugs targeting dyslipidemia. Maximizing the long-term safety of these drugs is important because they are often used preventatively and for prolonged periods by wide sectors of the population. Given this use, the long-term, population-level side effects of these drugs are especially important to minimize. Such an understanding could be achieved by investigating health impacts of genetic proxies for protein deficiencies that are associated with lipid levels. A study assessing this question exists for the chief lipoprotein(a)-modulating gene *LPA*[14]. However, it is less well known what are the health effects of the other major lipid-modifying genes targeted by drugs currently under development: *ANGPTL3, ANGPTL4, APOC3* and *CETP*[3-8]. Although some of these drug targets have undergone clinical trials, they do not consider long-term health impacts and suffer from confounding factors, such as compound-specific off-target effects.

Considering these clinical needs, the present study aimed to further the treatment of dyslipidemia in two ways. The primary goal was to identify genetic mechanisms to find new and safe drug targets for treating dyslipidemia and cardiometabolic diseases. The secondary aim was to evaluate the long-term health consequences of existing dyslipidemia drug targets. Studying Finns provides a promising avenue for reaching both of these goals. The Finnish population isolate shows an enrichment of protein-truncating variants (PTVs)[15, 16] (stop-gained, frame-shift and essential splice-site mutations), thus enabling the detection of both new and previously-known therapeutic effects through a smaller sample size than in the non-Finnish European population. Thanks to national health records on Finns, it is also possible to screen for a wide range of long-term health impacts associated with these PTVs. When combined, the data on PTVs and health records provide us with exactly the type of long-term, population-wide, on-target side effect data that is currently lacking, as discussed above.

## Results

An overview of our study’s analyses and their results is presented in Fig. 1. Our results are divided into three parts: the primary, secondary and tertiary analyses. Results of the primary analysis concern associations between PTVs and serum lipid levels. Results of the secondary analysis concern associations between lipid-associated PTVs and cardiometabolic disease risk. Finally, results of the tertiary analyses concern lipid and cardiometabolic disease-associated PTVs and their potential side effects.

**Figure 1.**
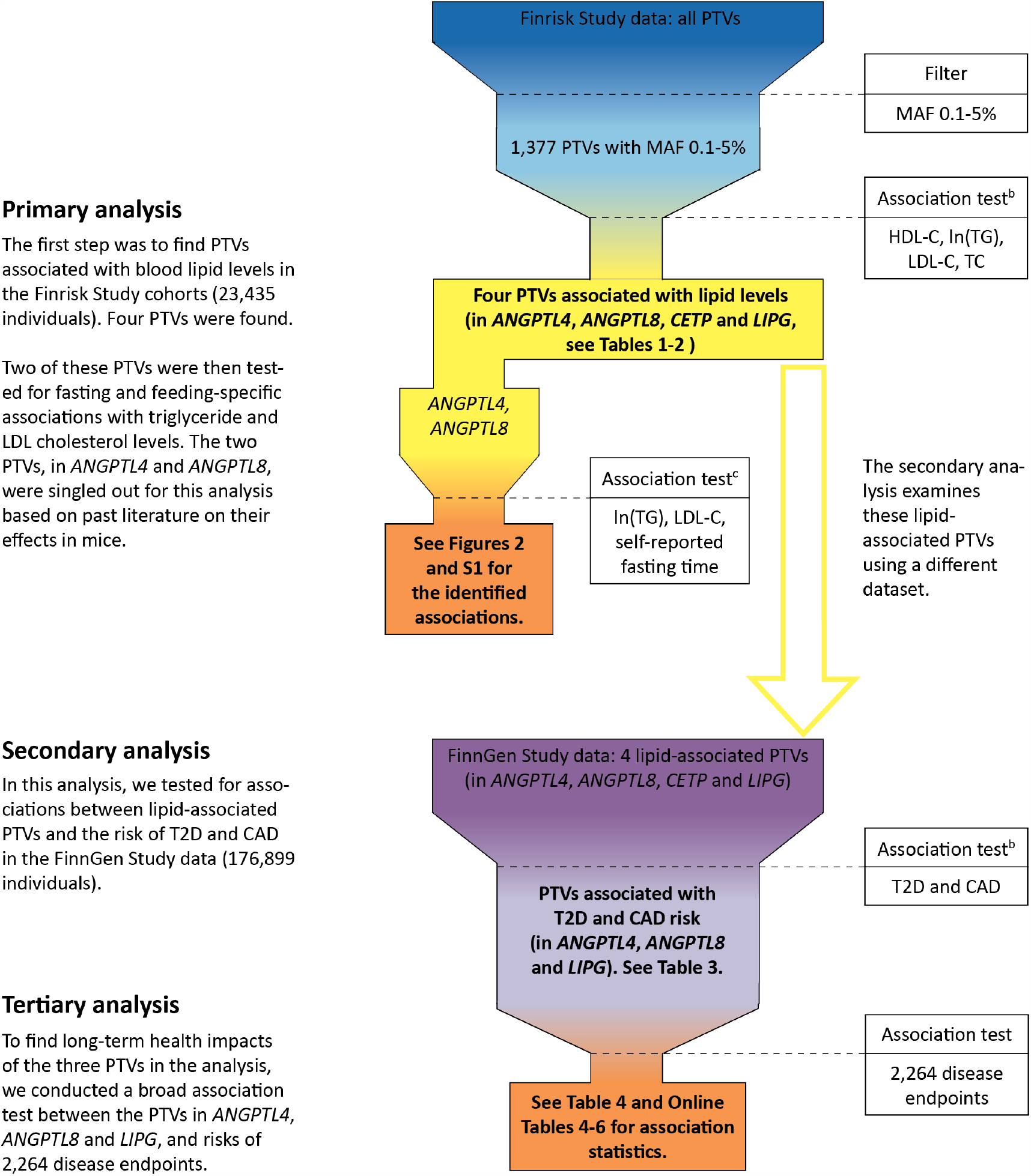
An overview of the present study.^a^. Abbreviations: PTV, protein-truncating variant; MAF, minor-allele frequency; HDL-C, high-density lipoprotein cholesterol level; ln(TG), natural logarithm of triglyceride level; LDL-C, low-density lipoprotein cholesterol level; TC, total cholesterol level; T2D, type 2 diabetes; CAD, coronary artery disease. ^a^ Shown in the figure is an overview of the present study. The final results are shown in boxes with a bolded font. ^b^ Results for these association tests were also compared with other PTVs in UK Biobank. ^c^ These association tests were post-hoc in nature.

### Primary analyses

#### Protein-truncating variants and serum lipid levels

We found four PTVs to be associated with serum lipid levels. PTVs in *CETP* (rs751916721-T), *LIPG* (rs200435657-A), and *ANGPTL8* (rs760351239-T) showed genome-wide significant associations with HDL cholesterol levels. PTVs in *ANGPTL4* (rs746226153-G) and *ANGPTL8* (rs760351239-T) had genome-wide significant associations with triglyceride levels (Tables 1-2). The four PTVs showed 27 to 210-fold enrichment in Finns compared to non-Finnish Europeans in the gnomAD database[17], version 2.1.1 (gnomad.broadinstitute.org) (Table 1).

**Table 1.** Associations between protein-truncating variants and serum lipid levels in the Finrisk Study.^a^.

| Locus and variant | Chromosome position<sup>b</sup> | Type of mutation | MAF in Finrisk Study % | Enrichment<sup>c</sup> | Primary association | *P* | Conditional *P*<sup>d</sup> |
| --- | --- | --- | --- | --- | --- | --- | --- |
| *ANGPTL4*<br>rs746226153-G | 19:8364556 | Frameshift | 0.48 | 27.2 | Triglycerides | $4.3 \times 10^{-9}$ | $3.7 \times 10^{-8}$ |
| *ANGPTL8*<br>rs760351239-T | 19:11240228 | Stop-gained | 0.15 | 85.0 | Triglycerides | $3.4 \times 10^{-9}$ | - |
| *CETP*<br>rs751916721-T | 16:56962097 | Splice-site | 0.12 | 46.3 | HDL cholesterol | $1.6 \times 10^{-21}$ | $5.9 \times 10^{-19}$ |
| *LIPG*<br>rs200435657-A | 18:49565316 | Splice-site | 0.20 | 210.1 | HDL cholesterol | $5.0 \times 10^{-13}$ | $3.6 \times 10^{-12}$ |

Abbreviations: MAF, minor-allele frequency.
<sup>a</sup> Shown in the table are the four PTVs associated genome-wide significantly (two-sided $P < 5 \times 10^{-8}$ ) with at least one serum lipid level measure in the Finrisk Study cohorts. The tested serum lipid levels were LDL, HDL and total cholesterol, as well as natural logarithm transformed triglycerides.
<sup>b</sup> Chromosome numbers and positions refer to genome build GRCh38.
<sup>c</sup> The allele frequency enrichment in Finns with respect to non-Finnish Europeans according to the gnomAD database, version 2.1.1 (gnomad.broadinstitute.org).
<sup>d</sup> The conditional P value is the largest P value from association tests conditioning on other previously reported genome-wide significant markers in the same gene. The conditional P value for rs760351239-T is not available because no genome-wide significant genetic variant in *ANGPTL8* has been reported before.

To evaluate whether the associations between the serum lipid levels and the four PTVs were independent, we conducted conditional tests of independence and determined which credible sets the PTVs belonged to. In our conditional analysis, the associations between lipid levels and the PTVs in the Finrisk Study[18] data were not explained by previously-reported genome-wide significant variants[19-22] (Table 1 and Tables S3-S6 in the Supplement). The *LIPG* and *ANGPTL8* PTVs alone formed credible sets with posterior probabilities above 99.9%. The *CETP* and *ANGPTL4* PTVs were in high linkage disequilibrium with the noncoding variants rs566571297-T (2^3^ = 0.99) and rs919624228-G (2^3^ = 0.97) respectively. With their correlated variant pair, these two PTVs formed credible sets with posterior probabilities higher than 99%. Hence, all the four PTVs were independently associated with changes in lipid levels with a very high probability (Tables S7-S10 in the Supplement).

To inspect if the lipid level associations with PTVs in Finns were concordant with other PTVs in the same genes, we surveyed the UK Biobank data. Using another rare non-Finnish European-enriched *ANGPTL8* PTV (rs145464906-T) in Britons, we observed comparable HDL cholesterol and triglyceride level associations as with the PTV found in Finns (Table 2). PTVs in *ANGPTL4, CETP* or *LIPG* were not present in the UK Biobank data and thus we were not able to investigate the lipid level associations of other protein-truncating variation in these genes.

**Table 2.**
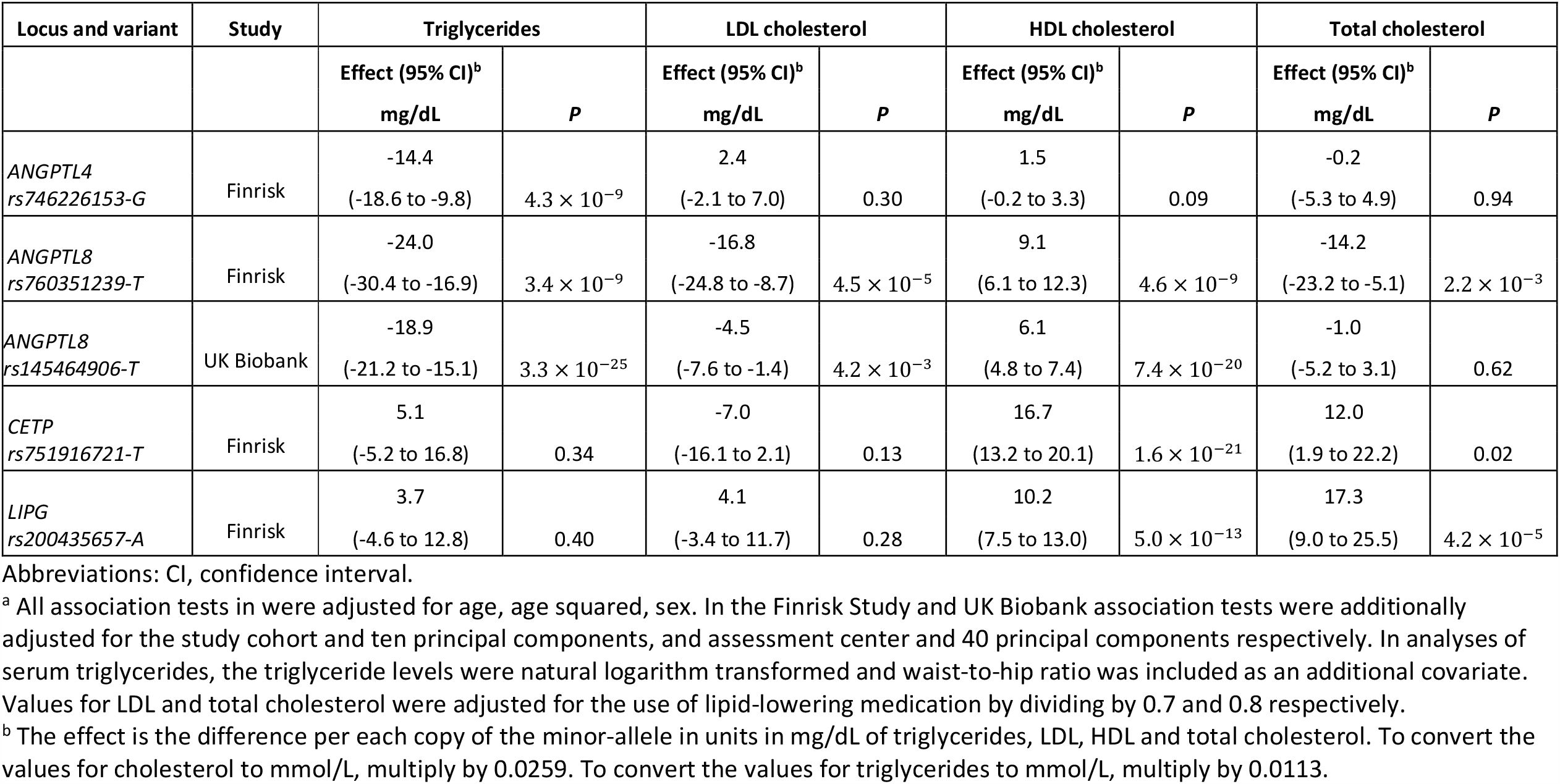
Associations between protein-truncating variants and serum lipid levels in the Finrisk Study and UK Biobank.^a^.

#### Protein-truncating variants and serum lipid levels as a function of fasting time

Next, we tested two hypotheses relating to the association of *ANGPTL4* and *ANGPTL8* with lipid levels as a function of fasting time. The first hypothesis was that *ANGPTL4* and *ANGPTL8* are associated with lower fasting and postprandial triglyceride levels respectively. We found that *ANGPTL4* PTV heterozygotes had 16.6% (20.2 mg/dL, *P* = 8.7 × 10^−7^) lower triglyceride levels than non-carriers when the time since the last meal was from 4 to 8 hours. On the other hand, the heterozygotes’ triglyceride levels were not significantly lower than non-carriers’ when the fasting time was 3 hours or less (14.8% higher [20.6 mg/dL], *P* = 0.09). The *ANGPTL8* PTV heterozygotes in turn had 34.3% (47.8 mg/dL, *P* = 0.01) lower triglyceride levels than non-carriers when the fasting time was up to 3 hours. From 4 to 8 hours after the last meal, the carriers had only 22.9% (27.9 mg/dL, *P* = 3.2 × 10 ^−7^) lower triglyceride levels than noncarriers.

The second hypothesis was that the *ANGPTL8* PTV is associated with lower fasting LDL cholesterol and triglyceride levels. We observed that *ANGPTL8* PTV heterozygotes had 13.4% (18.0 mg/dL, *P* = 4.5 × 10^−5^) lower LDL cholesterol levels than noncarriers with a fasting time between 4 and 8 hours. Moreover, the heterozygotes had 48.6% (55.1 mg/dL, *P* = 9.7 × 10^−5^) and 26.2% (36.9 mg/dL, *P* = 0.007) lower triglyceride and LDL cholesterol levels respectively than noncarriers when fasting for 9 hours or longer. The triglyceride and LDL cholesterol levels as a function of *ANGPTL4* and *ANGPTL8* PTV genotype and fasting time are shown in Fig. 2 and Fig. S1 in the Supplement respectively.

**Figure 2.**
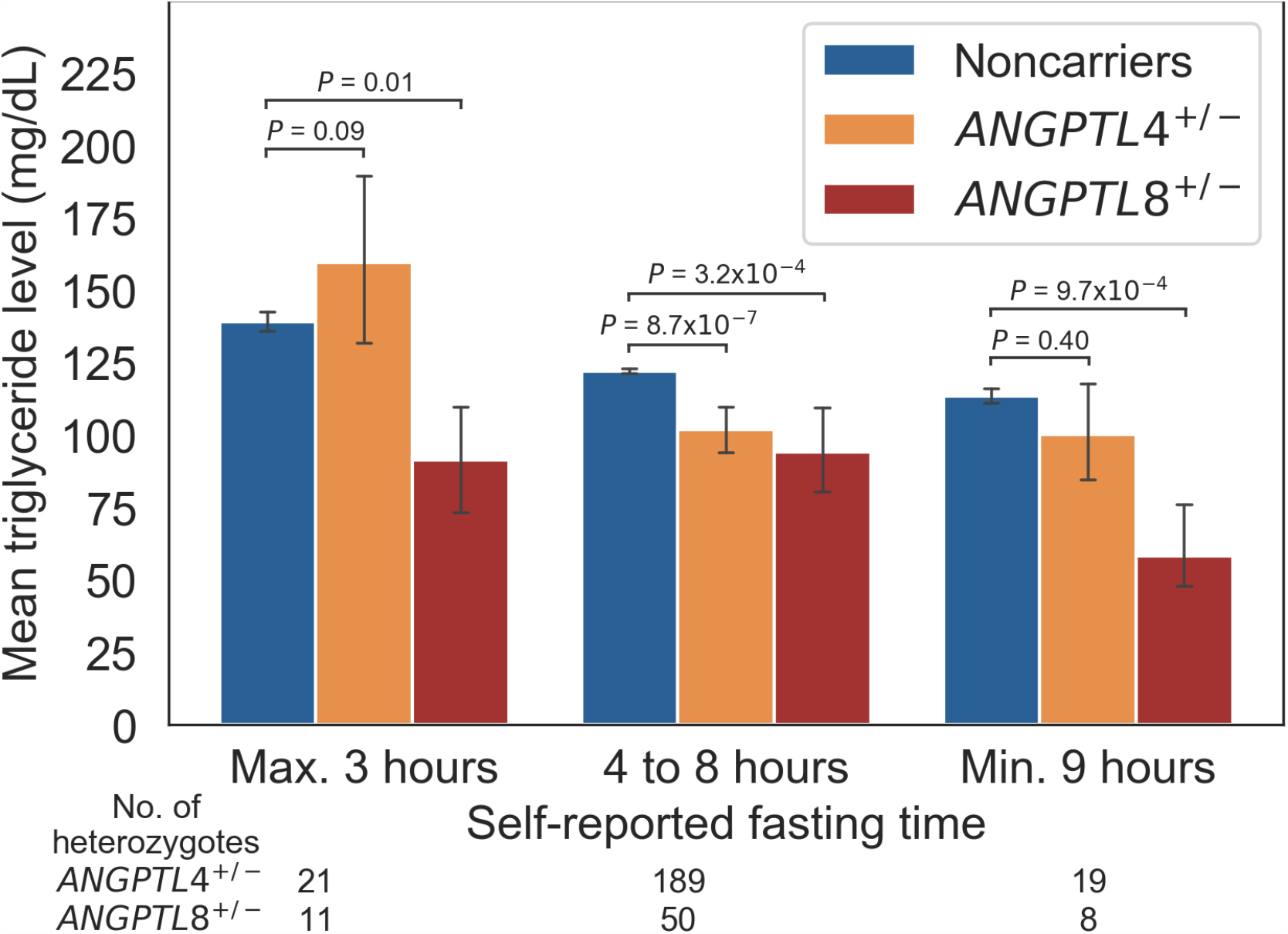
Mean serum triglyceride levels of *ANGPTL4* and *ANGPTL8* PTV heterozygotes at their self-reported fasting time in the Finrisk Study.^a^. ^a^ The figure shows the mean serum triglyceride level by *ANGPTL4* and *ANGPTL8* PTV carrier status with respect to self-reported fasting time. The number of *ANGPTL4* and *ANGPTL8* heterozygotes for each fasting time interval are reported below the fasting time legend. The height of the bars indicate means and the error bars 95% confidence intervals. The P values are for the two-sided Welch’s t-test of the natural logarithm of triglyceride levels between noncarriers and heterozygotes.

### Secondary analyses

#### Lipid-associated protein-truncating variants and cardiometabolic disease risk

Given that the primary analysis suggested that *ANGPTL4* and *ANGPTL8* PTVs may reduce triglyceride levels, we tested these PTVs’ impact on type 2 diabetes (T2D), coronary artery disease (CAD) risk in 176,899 individuals from the FinnGen Study. The *ANGPTL8* PTV was associated (two-tailed *P* < 0.05) with lower odds of CAD (OR = 0.56[0.38-0.83], *P* = 0.004) and T2D (OR = 0.65[0.47-0.92], *P* = 0.01). The *ANGPTL4* PTV heterozygotes had 30% lower odds of T2D (OR = 0.70[0.60-0.82], *P* = 1.4× 10^−5^). In tests between cardiometabolic diseases and the HDL cholesterol-increasing PTVs, we observed that the *LIPG* PTV was associated with 33% lower odds of T2D (OR = 0.67[0.48-0.91], *P* = 0.01). Finally, we tested if the four PTVs identified in our primary analysis were associated with available traditional non-lipid risk factors for CAD in the FinnGen Study (Table S11 in the Supplement).

Finally, as in the primary analysis, we checked if our findings for the Finnish-enriched *ANGPTL8* PTV (rs760351239-T) were congruent with the other *ANGPTL8* PTV enriched in UK Biobank. In specific, we tested if the non-Finnish European-enriched *ANGPTL8* PTV (rs145464906-T) was associated with T2D and CAD risk. Although rs145464906-T in the UK Biobank data alone was not significantly associated with the risk of T2D or CAD, a meta-analysis of the rs760351239-T and rs145464906-T PTVs strengthened both of these associations (Table 3).

**Table 3.** Association between protein-truncating variants and cardiometabolic disease risk in the FinnGen Study and UK Biobank.^a^.

| Locus and variant | Cohort | Type 2 diabetes; 23,338 cases in FinnGen |  |  |  | Coronary artery disease; 18,295 cases in FinnGen |  |  |  |
| --- | --- | --- | --- | --- | --- | --- | --- | --- | --- |
|  |  | Allele frequency<sup>b</sup> |  | OR<sup>c</sup><br>(95% CI) | *P* | Allele frequency<sup>b</sup> |  | OR<sup>c</sup><br>(95% CI) | *P* |
|  |  | Cases<br>% | Controls<br>% |  |  | Cases<br>% | Controls<br>% |  |  |
| *ANGPTL4*<br>*rs746226153-G* | FinnGen | 0.42 | 0.54 | 0.70<br>(0.60-0.82) | $1.4 \times 10^{-5}$ | 0.52 | 0.54 | 1.00<br>(0.83-1.21) | 1.00 |
| *ANGPTL8*<br>*rs760351239-T* | FinnGen | 0.09 | 0.12 | 0.65<br>(0.47-0.92) | 0.01 | 0.09 | 0.12 | 0.56<br>(0.38-0.83) | 0.004 |
| *ANGPTL8*<br>*rs145464906-T* | UK Biobank | 0.04 | 0.05 | 0.79<br>(0.48-1.30) | 0.35 | 0.03 | 0.05 | 0.73<br>(0.44-1.19) | 0.21 |
| *ANGPTL8* PTV<br>meta-analysis<sup>d</sup> | FinnGen and<br>UK Biobank | 0.06 | 0.07 | 0.69<br>(0.52-0.92) | 0.01 | 0.05 | 0.07 | 0.62<br>(0.45-0.84) | 0.002 |
| *CETP*<br>*rs751916721-T* | FinnGen | 0.10 | 0.011 | 0.82<br>(0.58-1.16) | 0.26 | 0.12 | 0.11 | 0.93<br>(0.62-1.37) | 0.70 |
| *LIPG*<br>*rs200435657-A* | FinnGen | 0.10 | 0.14 | 0.67<br>(0.48-0.91) | 0.01 | 0.13 | 0.13 | 0.71<br>(0.50-1.02) | 0.06 |

Abbreviations: OR, odds ratio; CI, confidence interval.
<sup>a</sup> Shown in the table are the association statistics between the serum lipid-associated PTVs and type 2 diabetes and coronary artery disease risk.
<sup>b</sup> The allele frequencies are reported in percent. The rs145464906-T allele frequencies in cases and controls were calculated in 343,687 unrelated individuals in UK Biobank and are based on PheCodes[45] 250.2 and 411.4 for type 2 diabetes and coronary artery disease respectively.
<sup>c</sup> In the FinnGen data the odds ratios were calculated using SAIGE[51] saddle-point approximation-based score test after adjustment for age, sex, genotyping batch and ten principal components of ancestry. The odds ratios for the associations between the disease risks and PTVs in UK Biobank were obtained from Zhou et al.[51]
<sup>d</sup> The rs760351239-T and rs145464906-T PTVs present in FinnGen and UK Biobank genotype data respectively have been meta-analyzed.

### Tertiary analyses

#### Phenome-wide associations of protein-truncating variants in ANGPTL4, ANGPTL8 and LIPG

To evaluate a wide range of possible long-term health effects of the PTVs associated with both lipid levels and cardiometabolic disease risk, we tested the association between these PTVs and 2,264 curated disease endpoints. In addition to T2D risk, the *ANGPTL4* PTV was phenome-wide significantly associated (two-tailed *P* < 2.2 × 10^−5^) with lower risk of insulin treatment for diabetes (OR = 0.70[0.60-0.81], *P* = 5.9 × 10^−6^), and with endocrine and metabolic diseases (OR = 0.78[0.70-0.87], *P* = 1.5 × 10^−5^). The *ANGPTL8* PTV was phenome-wide significantly associated with 47% lower odds of statin therapy (OR = 0.53[0.40-0.71], *P* = 1.2 × 10^−5^). However, the *ANGPTL8* PTV carriers had substantially higher risks of sensorineural hearing loss (OR = 2.45[1.63-3.68], *P* = 1.8 × 10^−5^) and esophagitis (OR = 174.3[17.7-1715.1], *P* = 9.7 × 10^−6^) (Table 4). See Online Tables 3-6 for the complete phenome-wide association statistics of the PTVs.

**Table 4.**
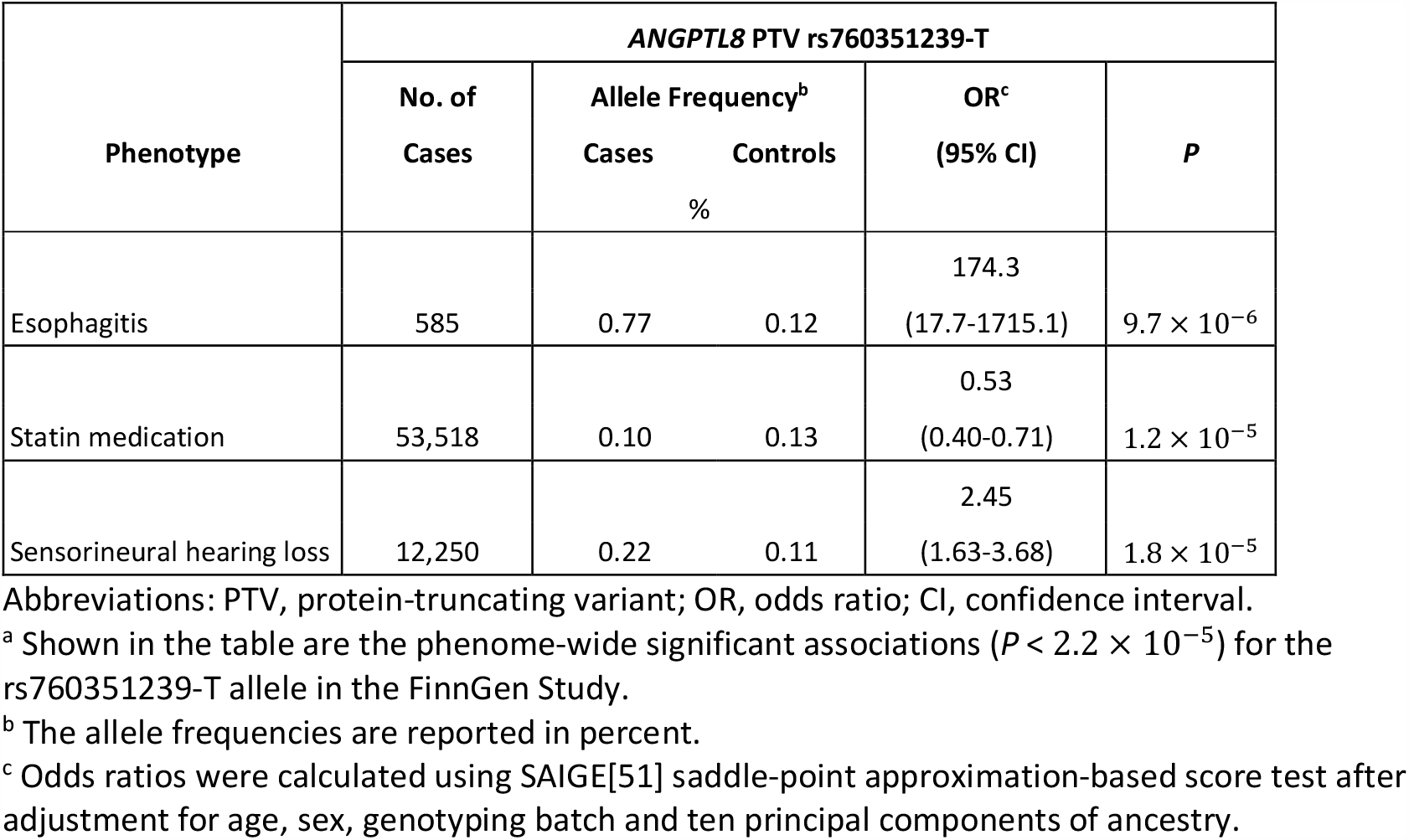
Phenome-wide associations with the *ANGPTL8* protein-truncating variant in the FinnGen Study.^a^.

## Discussion

In this study, we first presented associations between serum lipid levels and PTVs in four genes; *ANGPTL4, ANGPTL8, CETP* and *LIPG*. We then showed that carriers of PTVs in *ANGPTL8, ANGPTL4* and *LIPG* had lower risk for T2D and that the risk of CAD was lower in carriers of a PTV in *ANGPTL8*. Finally, we showed that the PTV carriers in *ANGPTL8* had elevated risk of esophagitis and sensorineural hearing loss.

First, our study points to *ANGPTL8* as a potential new therapeutic target for lowering triglyceride levels. This is interesting because while *CETP, LIPG* and *ANGPTL4* are well-known lipid genes, the association between a PTV in *ANGPTL8* and triglyceride levels has not previously been reported in humans. ANGPTL8 is known to inhibit triglyceride hydrolyzing lipoprotein lipase[23, 24]. In our data, the association between *ANGPTL8* PTV and lower triglyceride levels had the strongest effects soon after a meal in a post-prandial state, and after prolonged fasting. These findings are in line with animal studies which show that Angptl8 is secreted after feeding and has a very short half-life[25, 26], and that hepatic very low-density lipoprotein (VLDL) secretion is decreased in Angptl8 knockout mice[27].

Our results also show that carriers of a Finnish-enriched *ANGPTL8* PTV had 44% lower odds of coronary artery disease and 35% lower odds of type 2 diabetes. This is in line with genetic evidence of triglycerides being a causal factor for CAD risk[28]. Examining another non-Finnish European-enriched *ANGPTL8* PTV (rs145464906-T) in the UK Biobank data, we observed that also this PTV was associated with lower triglyceride levels. In addition, a meta-analysis of the *ANGPTL8* PTVs in FinnGen and UK Biobank data increased the statistical significance of T2D and CAD risk associations observed in FinnGen data alone.

Third, the *ANGPTL8* PTV carriers had an elevated risk of esophagitis and sensorineural hearing loss compared to noncarriers. Esophagitis is inflammation that damages the esophageal epithelium, which is usually a consequence of epithelial injury. When gastrointestinal reflux injures esophageal tissue, a pro-inflammatory response is caused by increased nuclear factor kappa-B kinase (NF-κB) mediated gene expression[29-31]. ANGPTL8 has been shown to affect NF-κB by suppressing the activation of its subunit gamma IKKγ[32] in humans. This mechanistic understanding is congruent with the observed high risk of esophagitis in *ANGPTL8* PTV carriers. We have no mechanistic explanation for the increased risk of sensorineural hearing loss which was associated with the *ANGPTL8* PTV, however this association was large enough to be of interest.

Thus, the increased risks of sensorineural hearing loss and esophagitis are findings with important implications for the safety of ANGPTL8 inhibition.

Lastly, in addition to this main contribution on the potential benefits and side effects of the *ANGPTL8* PTV, our study also found associations between the *ANGPTL4* PTV, the *LIPG* PTV and a lower risk of T2D. An earlier report also found that *ANGPTL4* PTV carriers had a lower risk of T2D than noncarriers[33]. The association between the *LIPG* PTV and a lower T2D risk supports previous findings showing that endothelial lipase, encoded by *LIPG*, is correlated with T2D risk[34, 35] and has pro-inflammatory properties[36-38]. Given that inflammation in adipose tissue increases insulin resistance[39-41], it may be expected that endothelial lipase affects T2D risk.

Our study has also some limitations. First, our study was observational in nature and consequently unable to directly reveal causal effects between genes and outcomes. Although we found robust lipid level associations for PTVs that were congruent with previous reports, we cannot confirm the causality between the PTVs and the observed disease risk associations. To confirm the causality between the associated disease risks and the Finnish-enriched PTVs, our findings need to be studied using human cell lines. However, utilizing samples from the bottlenecked Finnish population offered us considerable boosts in statistical power and testing these variants in non-bottlenecked populations would have only been possible with one or two orders of magnitude larges sample sizes.

In summary, we identified PTVs in *ANGPTL4* and *ANGPTL8* that were associated with lower triglyceride levels and PTVs in *CETP* and *LIPG* that were associated with higher HDL cholesterol levels. The carriers of PTVs in *ANGPTL8, ANGPTL4* and *LIPG* had lower risk of T2D and that the risk of CAD was lower in carriers of a PTV in *ANGPTL8*. Finally, we showed that the PTV carriers in *ANGPTL8* had elevated risk of esophagitis and sensorineural hearing loss than noncarriers. These findings point to potential target genes for development of novel preventive medication against T2D and CAD and highlight the utility of bottleneck populations in search of associations between protein-truncating variation and biomarkers.

## Methods

We identified PTVs associated with both lipid levels and cardiometabolic disease risk, and then examined their associations with other disease endpoints. In our primary analysis, we studied 23,435 Finns to find PTVs associated with serum lipid levels. Using data from the FinnGen Study, which is based on 176,899 individuals, we then studied the association between these mutations and risk of T2D and CAD. We refer to this as our secondary analysis. In our tertiary and final analysis, using FinnGen data, we assessed the long-term health impacts of the PTVs with cardiometabolic disease risk associations by screening them for modified risk of 2,264 diseases. An overview of our study is depicted in Fig. 1.

### Study populations

We used three different data sets for our study: the Finrisk Study cohorts, the FinnGen Study and UK Biobank. In total, the Finrisk Study dataset contained 23,435 chip-genotyped and imputed samples, selected randomly from the Finnish population in 1992, 1997, 2002, 2007 and 2012[18]. The baseline characteristics of the Finrisk Study participants are shown in Table S1 in the Supplement.

The FinnGen Study contains biobank data and national health registry data for 176,899 individuals. The health registry information of participants from the Finrisk and FinnGen Study was followed up until 31.12.2017. The details of the individual FinnGen cohorts are shown in Table S2 in the Supplement. All Finrisk and FinnGen Study participants were of Finnish descent. The genotyping and imputation data release of UK Biobank data was from 5th March 2018.

All participants gave written informed study-specific consent. Patients and control subjects in the FinnGen Study provided informed consent for biobank research, based on the Finnish Biobank Act. Alternatively, older Finnish research cohorts, collected prior the start of FinnGen Study (August 2017), were collected based on study-specific consents and later transferred to the Finnish biobanks after approval by Fimea, the National Supervisory Authority for Welfare and Health. Recruitment protocols followed the biobank protocols approved by Fimea. The Coordinating Ethics Committee of the Hospital District of Helsinki and Uusimaa (HUS) approved the FinnGen Study protocol No. HUS/990/2017.

The FinnGen project is approved by Finnish Institute for Health and Welfare (THL), approval number THL/2031/6.02.00/2017, amendments THL/1101/5.05.00/2017, THL/341/6.02.00/2018, THL/2222/6.02.00/2018, THL/283/6.02.00/2019), Digital and population data service agency VRK43431/2017-3, VRK/6909/2018-3, the Social Insurance Institution (KELA) KELA 58/522/2017, KELA 131/522/2018, KELA 70/522/2019 and Statistics Finland TK-53-1041-17.

The Biobank Access Decisions for FinnGen Study samples and data utilized in FinnGen Data Freeze 4 include: THL Biobank BB2017_55, BB2017_111, BB2018_19, BB_2018_34, BB_2018_67, BB2018_71, BB2019_7 Finnish Red Cross Blood Service Biobank 7.12.2017, Helsinki Biobank HUS/359/2017, Auria Biobank AB17-5154, Biobank Borealis of Northern Finland_2017_1013, Biobank of Eastern Finland 1186/2018, Finnish Clinical Biobank Tampere MH0004, Central Finland Biobank 1-2017, and Terveystalo Biobank STB 2018001.

### Genotyping and quality control

The Finrisk Study samples were genotyped using the HumanCoreExome BeadChip, Human610-Quad BeadChip, Affymetrix6.0 and Infinium HumanOmniExpress (Illumina Inc., San Diego, CA, USA) chips and a Finnish-ancestry-specific imputation panel consisting of 2,690 deep-coverage (25-30x) whole-genome and 5,092 whole-exome sequences. In the primary analysis, the 1,377 PTVs were located in 1,209 genes and had a MAF between 0.1 and 5%. These PTVs in the Finrisk Study cohorts were imputed and had a IMPUTE2[42] genotype information score with a mean of 0.95 (standard deviation of 0.05) and a minimum of 0.75. The FinnGen Study samples were genotyped with various Illumina and a custom AxiomGT1 Affymetrix array (www.finngen.fi/en/researchers/genotyping). All the lipid-associated PTVs were directly genotyped in at least 62.9% of the FinnGen Study individuals with the AxiomGT1 Affymetrix array. The genotypes of the PTVs in our secondary and tertiary analyses that were not genotyped on chip were imputed using a genotype panel that consisted of 3,775 deep-coverage (25-30x) whole-genome sequenced individuals of Finnish ancestry. The PTVs in our secondary and tertiary analyses had IMPUTE2[42] genotype information scores above 0.92 in the FinnGen data. Detailed description of the genotyping methods, genotype imputation and quality-control procedures are described in the Supplement.

### Study outcomes

A blood sample and the self-reported fasting time since the previous meal at the time of blood sample collection of each Finrisk Study participant were collected during a clinical visit. The total cholesterol, HDL cholesterol and triglyceride levels were measured directly from serum or plasma and LDL cholesterol was either directly measured or estimated using the Friedewald formula[43]. In UK Biobank the blood lipid levels were measured from serum directly. LDL and total cholesterol levels of individuals with lipid-lowering therapy were divided by 0.7 and 0.8[44] respectively.

Information on diagnoses in the FinnGen data were collected and confirmed by examining national healthcare registries and recorded using the *International Classification of Diseases* [ICD] revisions 8-10. Purchase information on prescription drugs since 1995 were obtained from the Finnish social insurance institution (KELA) reimbursement records and coded using the *Anatomical Therapeutic Chemical* [ATC] classification). All FinnGen Study participants’ healthcare registry information were followed up until 31.12 2017. Cancer diagnoses and causes of death were obtained from their respective national registries. The clinical expert groups of the FinnGen Study have defined disease events using ICD and ATC codes. For a complete list of the considered clinical endpoints and corresponding ICD and ATC codes, see Online Tables 1-3.

For the T2D and CAD statuses in UK Biobank participants we used PheCodes[45] 250.2 and 411.4 respectively.

### Study design and statistical analyses

#### Primary analysis

In this analysis, we sought to identify associations between lipid levels and PTVs in the Finrisk Study. The lipid levels tested were plasma or serum levels of HDL cholesterol, LDL cholesterol, total cholesterol and logarithmically transformed triglycerides on a natural log scale. Our model was additive, and included age, age squared, sex, collection year, and ten principal components of ancestry as fixed-effects covariates. To correct for the effect of adiposity on triglyceride levels, we adjusted the triglyceride association tests for waist-to-hip ratio as well. We considered the genome-wide association significance threshold of a two-sided P value of less than 5.0 × 10^−8^ to be significant. Genetic association analyses were carried out using the PLINK[46], version v1.90b3.45 (www.cog-genomics.org/plink/1.9/), file format, Python, version 3.6 (www.python.org) and the statsmodels Python package, version 0.8.0 (www.statsmodels.org). In our scan, we only considered variants with a MAF 0.1-5% to account for adequate statistical power and the expected low frequency of high-impact alleles. To assess the functional validity of the associations, we performed conditional analyses with previously associated variants in the gene (Tables S3-S6 in the Supplement) and determined the 95% credible sets of variants in each gene locus (Tables S7-S10 in the Supplement). The credible sets were determined using FINEMAP[47], version 1.4.3 (www.finemap.me).

Next, we carried out a post-hoc analysis to test the hypotheses that *ANGPTL4* and *ANGPTL8* PTVs are associated with triglycerides and LDL cholesterol levels as a function of fasting time. These hypotheses were based on animal studies that we reviewed after finding four PTVs associated with lipids in our genome-wide analysis. Firstly, we found studies that showed that in mice, Angptl4 and Angptl8 inhibit lipoprotein lipase (LPL) as a function of fasting time[25, 26, 48]. LPL inhibition is important because it is the mechanism by which several triglyceride-lowering drugs currently under development work[3, 4, 7]. If the association between triglyceride levels and *ANGPTL8* and *ANGPTL4* PTVs depends on fasting time, then fasting time dependent effects on triglyceride levels of these PTVs are very relevant for our study, which aims to assess the effect of these PTVs on hypertriglyceridemia risk.

Literature similarly suggested that, to determine whether *ANGPTL8* is a viable drug target, it might also be important to test its association with LDL cholesterol levels as a function of fasting time. A mouse study found that *ANGPTL8* modulates VLDL secretion[27]. Given our goal of testing the viability of *ANGPTL8* as a drug target, the effect of *ANGPTL8* on VLDL levels would be important to assess in humans because high VLDL levels are associated with a higher risk of CAD[49]. We could not directly test whether human *ANGPTL8* PTV carriers had lower VLDL levels than noncarriers, so we instead tested the association between the *ANGPTL8* PTV and levels of

LDL cholesterol and triglycerides after fasting. Our rationale for this test was that *fasting* triglyceride and LDL cholesterol levels in particular can be considered a proxy for VLDL. This is due to the deficiency of chylomicron and chylomicron remnant particles in the blood stream in a fasted state[50].

We examined the serum lipid level associations of other PTVs in the same genes as in our findings, using UK Biobank data. For this we used another rare non-Finnish European-enriched *ANGPTL8* PTV (rs145464906-T). PTVs in *ANGPTL4, CETP* or *LIPG* were not present in the UK Biobank data and thus we were not able to analyze the lipid level associations of protein-truncating variation in these genes.

#### Secondary and tertiary analyses

In the secondary analyses, we examined associations between cardiometabolic disease risk and the lipid-associated PTVs identified in the primary analysis. The analyses included two cardiometabolic diseases: T2D and CAD. The associations (two-tailed *P* < 0.05) were tested on data from the FinnGen Study.

As in the primary analysis, we checked if our findings for the Finnish-enriched *ANGPTL8* PTV (rs760351239-T) were consistent with another rare *ANGPTL8* PTV in UK Biobank. We tested if the non-Finnish European-enriched *ANGPTL8* PTV (rs145464906-T) was associated with T2D and CAD risk. In addition, we conducted an inverse-variance-weighted meta-analysis of the rs760351239-T and rs145464906-T PTVs in the FinnGen and UK Biobank data.

In the tertiary analyses we examined the other health impacts of the PTVs associated with both serum lipid levels and the risk of T2D or CAD. We screened these PTVs broadly for modified risk of 2,264 diseases in the FinnGen data. We regarded a two-sided P value below 2.2 × 10^−5^ (Bonferroni-corrected threshold for 2,264 traits) to be statistically significant.

In both the secondary and tertiary analyses in the FinnGen data, the odds ratios for disease outcomes were estimated using SAIGE[51], version 0.35.8.8 (www.github.com/weizhouUMICH/SAIGE/releases/tag/0.35.8.8). Age, sex, genotyping batch and ten principal components of ancestry and the kinship matrix were included as fixed-effects covariates. See Online Tables 1-3 for the disease definitions and Online Tables 4-6 for the associations between disease risks and the PTVs. The details of computing the associations in the FinnGen data are described in the Supplement. The T2D and CAD risk associations were also computed using SAIGE and were obtained from Zhou et al.[51]

## Data Availability

The FinnGen data may be accessed through the FinnBB portal (www.finbb.fi) of the Finnish Biobanks and THL Biobank data through THL Biobank (https://thl.fi/en/web/thl-biobank).

https://thl.fi/en/web/thl-biobank

## Acknowledgements

We would like to thank Heidi Silvennoinen and Katri Silvennoinen for proofreading the manuscript, and Sari Kivikko, Huei-Yi Shen and Ulla Tuomainen for management assistance. Many thanks go to Kalle Pärn, Marita A. Isokallio, Javier Nunez Fontarnau and Priit Palta for the genotype imputation of Finrisk Study participants.

Following biobanks are acknowledged for collecting the FinnGen project samples: Auria Biobank (https://www.auria.fi/biopankki), THL Biobank (https://thl.fi/fi/web/thl-biopankki), Helsinki Biobank (https://www.terveyskyla.fi/helsinginbiopankki), Biobank Borealis of Northern Finland (https://www.oulu.fi/university/node/38474), Finnish Clinical Biobank Tampere (https://www.tays.fi/enUS/Research_and_development/Finnish_Clinical_Biobank_Tampere), Biobank of Eastern Finland (https://itasuomenbiopankki.fi), Central Finland Biobank (https://www.ksshp.fi/fi-FI/Potilaalle/Biopankki), Finnish Red Cross Blood Service Biobank (https://www.veripalvelu.fi/verenluovutus/biopankkitoiminta) and Terveystalo Biobank (https://www.terveystalo.com/fi/Yritystietoa/Terveystalo-Biopankki/Biopankki/). All Finnish Biobanks are members of BBMRI.fi infrastructure (www.bbmri.fi). We also thank study participants for their generous participation at THL Biobank, the National FINRISK study and UK Biobank. This research has been conducted using the UK Biobank Resource with the application number 22627.

The FinnGen project is funded by two grants from Business Finland (HUS 4685/31/2016 and UH 4386/31/2016) and by eleven industry partners (AbbVie Inc, AstraZeneca UK Ltd, Biogen MA Inc, Celgene Corporation, Celgene International II Sàrl, Genentech Inc, Merck Sharp & Dohme Corp, Pfizer Inc., GlaxoSmithKline, Sanofi, Maze Therapeutics Inc., Janssen Biotech Inc).

This work was supported by the Sigrid Jusélius Foundation (to S.R., A.P.); University of Helsinki HiLIFE Fellow grants 2017-2020 (to S.R.); Academy of Finland Center of Excellence in Complex Disease Genetics (grant number 312062 to S.R., 312074 to A.P., 312075 to M.J.D.); Academy of Finland (grant number 285380 to S.R., 128650 to A.P., 321356 to A.S.H.); Foundation and the Horizon 2020 Research and Innovation Programme (grant number 667301 [COSYN] to A.P.); The Finnish Foundation for Cardiovascular Research (to S.R., A.P., V.S.); Precision Health Scholars Award from the University of Michigan Medical School (to I.S.); Doctoral Programme in Population Health, University of Helsinki (to P.H., T.K.); Emil Aaltonen Foundation (to P.H.). The funders had no role in the study design, data collection and analysis, decision to publish or preparation of the manuscript.

## Data availability

The FinnGen data may be accessed through Finnish Biobanks’ FinnBB portal (www.finbb.fi) and THL Biobank data through THL Biobank (https://thl.fi/en/web/thl-biobank).

